# Development and Evaluation of an ARTIC-Based Amplicon Sequencing Assay for Whole-Genome Characterization of Respiratory Syncytial Virus

**DOI:** 10.64898/2026.04.06.26350258

**Authors:** Keri Smith, James Martinez, Harrison Yu, Jason Harrison, Caleb Umunna, Blake Bertrand, Michael Heck, Ellen N. Kersh, Nandhakumar Balakrishnan, Tonia Parrott, Arunachalam Ramaiah

**Author notes:** Corresponding author Arunachalam Ramaiah, MS, PhD.

## Abstract

Respiratory syncytial virus (RSV), a ∼15.2 kb negative-sense RNA virus, causes acute respiratory infections in infants and older adults. Its two subtypes, RSV-A and RSV-B, evolve rapidly, making ongoing monitoring of circulating strains essential. The Georgia Public Health Laboratory (GPHL) developed and evaluated an amplicon-based whole-genome sequencing (WGS) assay for RSV surveillance. A total of 214 de-identified remnant clinical specimens (102 RSV-A; 112 RSV-B) with RT-PCR Ct values <31 were included. RSV genomes were amplified using ARTIC-style and custom primer sets, with the ARTIC set showing superior performance. Libraries were prepared using a modified Illumina COVIDSeq protocol, sequenced on NextSeq 1000/2000 instruments, and analyzed using the GPHL-RSV-PIPE bioinformatics pipeline. Among genomes meeting validation criteria, sequencing depth was slightly higher for RSV-A (median 53,433×; mean 51,076×) than RSV-B (median 49,699×; mean 46,945×), whereas genomic coverage was slightly lower for RSV-A (median 97.5%; mean 96.6%) than RSV-B (median 98.3%; mean 97.6%). Predominant lineages were A.D.3.1 and A.D.5.2 for RSV-A and B.D.E.1 for RSV-B. For RSV-A, the assay showed 92.8% accuracy, 96.2% sensitivity, 87.2% specificity, 92.6% positive predictive value, and 93.2% negative predictive value. Intra- and inter-run precision assessed using 16 and 53-57 genomes, respectively, showed nearly 100% consensus genome identity with 0–5 nucleotide differences. Specificity testing of 31 non-RSV specimens produced no false-positive detections. Limits of detection were 4.4 TCID_₅₀_/mL for RSV-A and 18.6 TCID_₅₀_/mL for RSV-B. These results demonstrate that the ARTIC-based RSV WGS assay enables near real-time surveillance and strengthens data-driven public health responses to future outbreaks.

**IMPORTANCE STATEMENT:** Respiratory syncytial virus (RSV), with two major subtypes, RSV-A and RSV-B, causes acute respiratory infections that can be severe in infants under six months and older adults. Current RSV surveillance at the Georgia Public Health Laboratory (GPHL) relies on the Thermo Fisher TaqMan™ Gene Expression Capillary assay, which detects and subtypes RSV but lacks resolution for lineage classification and identification of emerging variants. To address this critical gap, GPHL developed and evaluated an amplicon-based whole-genome sequencing (WGS) assay using 214 de-identified RSV clinical specimens. Genomes were amplified using ARTIC-style and custom-primer sets, with ARTIC primers showing superior performance. The assay demonstrated strong sequencing depth, genomic coverage, specificity, repeatability, reproducibility, and low limits of detection. RSV lineages were accurately determined based on genetic variation. These results establish that the ARTIC-based WGS assay enables near real-time genomic surveillance, supporting monitoring of circulating RSV strains and informing data-driven public health responses.

## INTRODUCTION

Human respiratory syncytial virus (RSV) is a seasonal, acute respiratory virus that primarily affects pediatric (under 5 years of age), geriatric (over 60 years of age), and immuno-compromised populations [1]. During the 2024-2025 season, RSV was associated with an estimated 190,000 to 350,000 hospitalizations, including severe cases of lower respiratory tract infections such as bronchiolitis and pneumonia [2–3]. Despite its relatively small genomic size of approximately 15.2 kilobases, about half the size of SARS-CoV-2, the RSV genome comprises ten genes encoding eleven proteins, with the fusion (F) and attachment glycoprotein (G) primarily serving as primary targets for vaccine development [4]. The two viral subgroups, RSV-A and RSV-B, commonly co-circulate and are classified based on antigenic variability in the G protein [5–6].

Recognition of RSV as a global health burden underscores the importance of tracking and surveilling associated outbreaks to support epidemiological efforts at the local, regional, and national levels. Current global pathogen genomic surveillance strategies emphasize expanding sustained capacity for threat detection to strengthen preparedness, readiness, response, and recovery cycle against future pandemics or epidemics [7]. Whole-genome sequencing (WGS) of RSV provides genome-wide resolution for each clinical specimen, enhancing existing public health surveillance through genomic characterization as well as clade and lineage assignment. These capabilities are enabled by next-generation sequencing (NGS) technologies, which sequence genomic material from patient clinical specimens and process it through bioinformatics workflows for genome assembly and downstream analyses. The resulting genomic data adds critical context to epidemiological investigations by enabling assessment of genetic relatedness among positive specimens, thus identifying potential transmission dynamics [8]. Beyond outbreak investigation, genomic surveillance supports the identification of emerging lineages that is essential for the development of seasonal vaccines and monoclonal antibody therapies, ultimately informing immunization campaigns and treatment strategies that reduce the overall public health burden of RSV [4].

In recent years, NGS efforts have become essential in public health laboratories and epidemiological studies for pathogen genomic surveillance and outbreak investigations [9–17]. The Georgia Department of Public Health (GDPH) has routinely conducted viral respiratory disease surveillance for RSV to monitor infections across the state of Georgia [18]. The Georgia Public Health Laboratory (GPHL) of GDPH currently employs the Thermo Fisher TaqMan™ Gene Expression Capillary assay, with the TrueMark™ Respiratory Panel 2.0, TaqMan™ Array card to detect and subtype RSV from nasopharyngeal swabs collected in viral transport medium (VTM). Partnerships between the GPHL, GDPH Emerging Infection Program (EIP) and clinical laboratories form the basis of this surveillance network, which reports total number of tests performed and laboratory-confirmed positives to the National Respiratory and Enteric Virus Surveillance System (NREVSS). While current surveillance methods such as RT-PCR or antigen testing are effective for case detection and tracking, they provide limited resolution for deeper genomic epidemiological analysis [18]. Recent expansion of NGS capacity at GPHL for pathogens of public health importance has enabled a transition towards genomic surveillance approaches. For RSV-A and RSV-B genomic surveillance, GPHL has developed a tiled amplicon-based methodology through modification of the Illumina COVIDSeq Test (RUO Version) kit. The methodology relies on overlapping primer pairs that span the RSV-A and RSV-B genomes exclusively that produce highly specific amplicons usable for detection and subtyping [16]. This study describes the development and validation of a WGS assay and an accompanying bioinformatics pipeline for real-time RSV genomic surveillance for circulating strain characterization. Implementation of this RSV workflow will expand viral respiratory pathogen sequencing capabilities at GPHL and provide a basis for the continued development of WGS-based genomic surveillance assays in other public health laboratories.

## METHODS

### Amplicon Panel Design and Evaluation

We initially evaluated the performance of two amplicon panels: a previously published ARTIC-style RSV primer set [16] and a custom primer set designed in-house. The custom primer set was developed using the PrimalScheme tool version 1.4.1 via command line interface [19]. For primer design, 82 high-quality RSV-A genomes and 101 RSV-B genomes retrieved from the GISAID database were aligned using Clustal Omega tool (web version 2.1) [20]. Based on percent identity matrix results, sequences with <95% or >99% identity were excluded. Following filtering, six RSV-A and six RSV-B sequences, along with the reference sequences used as input for PrimalScheme tool to design the custom primer set [21]. Primer design parameters included target insert overlap size and amplicon length range. The custom primer set yielded 27 primer pairs for RSV-A and 33 primer pairs for RSV-B. An insert overlap size of 75 nucleotides and amplicon length range of 750-800 bp for RSV-A and 650-700 bp for RSV-B resulted in a predicted genome coverage of approximately 98% for both subtypes. Primer specificity was evaluated using the NCBI BLAST tool [22].

The ARTIC RSV panel comprised of 50 primer pairs for each of RSV-A and RSV-B, producing approximately 400 bp amplicons that collectively covered the entire genome. Both the custom-designed primers and the ARTIC RSV primer set [16] were synthesized by Integrated DNA Technologies (IDT). For library preparation, RSV-A and RSV-B primer schemes were divided into two pools based on even and odd primer numbering, resulting in 100 µM primer pools containing exclusively either even or odd primers for each subtype. All primer pools were stored at −20 °C. Following side-by-side analysis of sequencing coverage obtained with both primer sets, the ARTIC RSV primer set was selected for subsequent validation.

### Specimen Collection and Nucleic Acid Extraction

We used remnant clinical specimens for WGS assay development and validation. These specimens, obtained from nasopharyngeal swabs collected in Georgia, were previously received by the GPHL for routine testing and diagnostics. They were collected between October 2023 and April 2025 and stored in viral transport media at -80°C. All specimens were de-identified, and all personal health information (PHI) was removed prior to processing.

The nucleic acid extraction method was validated and automated with the liquid-handling Chemagic 360 instrument (PerkinElmer, cat no 2024-0020). Using the Chemagic Viral DNA/RNA 300 Kit H96 (PerkinElmer, cat no CMG-1033-S), each well in a 96 deep-well plate consisted of 300 μL of specimen, 300 μL of Lysis Buffer, 5 μL of Internal Control (Thermo Fisher TaqMan™ Xeno Control, cat no A39179), 4 μL of Poly(A) RNA, and 10 μL of Proteinase K. The sample plate, along with a round-bottom plate with 150 μL of Magnetic Beads, a deep-well plate prefilled with 600 μL Elution Buffer, and three empty deep-well plates, was loaded onto the instrument. The extraction yielded a total nucleic acid elution volume of 60 μL per sample. Intact synthetic RNA transcripts of RSV-A and RSV-B (ZeptoMetrix: RSV-A Cat. No. NATRSVA-STQ, RSV-B Cat. No. NATFRC-ERC; ATCC: RSV-A Cat. No. VR-3418, RSV-B Cat. No. VR-1400) were run in duplicate as positive controls, while nuclease-free water served as a negative extraction control. Following extraction, both RSV-A and RSV-B subtypes were detected and subtyped using the validated Thermo Fisher TaqMan™ Gene Expression Capillary (Thermo Fisher, Cat. No. A49047) real-time PCR assay. Specimens underwent a pre-amplification step followed by amplification, and those with C_q_<31 were considered positive. A total of 102 RSV-A and 112 RSV-B specimens, with a C_q_ range of 7-30, were ultimately selected for the validation.

The lower limit of detection (LOD) for RSV-A and RSV-B was determined using serial dilutions of ATCC reference material (ATCC, Cat. No. VR-3418 and Cat. No. VR-1400) with known concentrations reported in Tissue Culture Infectious Dose 50 (TCID_50_/mL). Viral RNA was extracted directly from diluted samples without prior RT-qPCR quantification, and concentrations were reported as provided by the manufacturer. For RSV-A, dilutions ranged from 280,000 to 0.35 TCID_50_/mL, and for RSV-B, dilutions ranged from 89,000 to 0.89 TCID_50_/mL. Each dilution scheme was run in quadruplicate.

### Library Preparation and Whole-Genome Sequencing

The Illumina COVIDSeq Test protocol and kit (RUO Version) (cat no 20043475) was adapted from previous studies [11, 23] for library preparation using the pre-programmed liquid handler, Sciclone G3 (PerkinElmer, Cat. No. CLS152236). RNA extracts were first converted to complementary DNA (cDNA), which was then used for PCR amplification. Nuclease-free water, serving as the negative (no template) control, was added in duplicate to the sample plate. Instead of the COVIDSeq primers, the ARTIC RSV-A and RSV-B primers were mixed equimolarly into two pools and diluted to 10 uM. Each sample underwent reverse transcription to synthesize the first strand of cDNA. This reaction included 8.5 uL of extracted sample, 8.5 uL of EPH3, and 8 uL of First Strand cDNA Master Mix (composed of 9 uL of FSM and 1 uL of RVT). Following reverse transcription, PCR amplification was performed with a reaction mixture containing 5 uL of cDNA, 15 uL of IPM, 4.3 uL of primer mix, and 4.7 uL of nuclease-free water. PCR thermocycling conditions followed the COVIDSeq Test protocol: 98°C for 3 minutes, then 35 cycles of 98°C for 15 seconds and 63°C for 5 minutes, followed by a hold at 4°C. PCR products were subsequently processed through tagmentation using the IDT for Illumina-PCR Indexes Sets 1-4 (384 Indexes; Illumina, Cat. No. 20043137). Library cleanup and pooling were performed manually. Library concentration and fragment size were determined using the Qubit 3.0 Fluorometer (Invitrogen, Cat. No. Q33216) with the Qubit dsDNA High Sensitivity Assay kit (Invitrogen, cat no Q32854) and the Agilent D1000 TapeStation (Agilent Technologies). Based on these results, libraries were normalized to a final loading concentration of 1000 pM and spiked with 1-2% PhiX internal control. WGS was performed on Illumina NextSeq1000 and NextSeq2000 sequencers using NextSeq1000/2000 P2 200-cycle cartridges and flow cells.

### Bioinformatics Pipeline and Data Analysis

A custom reference-based bioinformatics pipeline, GPHL-RSV-PIPE, adapted from the ITER pipeline [24] (Figure 1; Table 1), was developed and optimized for RSV-A and RSV-B detection, subtyping, genome assembly and lineage classification. Paired-end raw reads in FASTQ format were pre-processed by adapter trimming using fastp (v0.23.4) [25], followed by quality control (QC) assessment before and after trimming with FastQC (v0.12.1) [26]. Human reads were removed using Kraken2 (v2.1.3) [27]. Processed reads were then analyzed using an updated version of fastv, a kmer detection tool for identifying positive hits on RSV-A and RSV-B [28]. A sample was considered positive, if it met thresholds of >=10% kmer coverage and mean read depth of >=500 reads. Reads were initially aligned to both reference genomes, RSV-A (hRSV/A/England/397/2017, GISAID EPI_ISL_412866) and RSV-B (hRSV/B/Australia/VIC-RCH056/2019, GISAID EPI_ISL_1653999), using BBMap (v39.16) [29] to determine the subtype with the highest number of mapped reads. Based on the BBMap’s results, the appropriate reference genome was selected for final alignment and BAM file generation using BWA-MEM [30]. Primer BED files specific to RSV-A and RSV-B were provided during alignment. The resulting BAM files were sorted indexed using SAMtools [31] and primer sequence were trimmed with iVar trim (v1.4.2) [32]. Consensus sequence generation and variant calling were performed using iVar, with a minimum base quality threshold of 20 and a minimum read depth of 10. Percent genomic coverage and mean sequencing read depth across the genome were calculated using mosdepth (v0.3.8) [33]. Nextclade Web (v3.18.0) was used for lineage classification [34]. MultiQC was used to aggregate QC metrics generated throughout the pipeline [35]. All plots and figures were produced in R (v4.5.2) [36] using the ggplot2 package [37].

**Figure 1.**
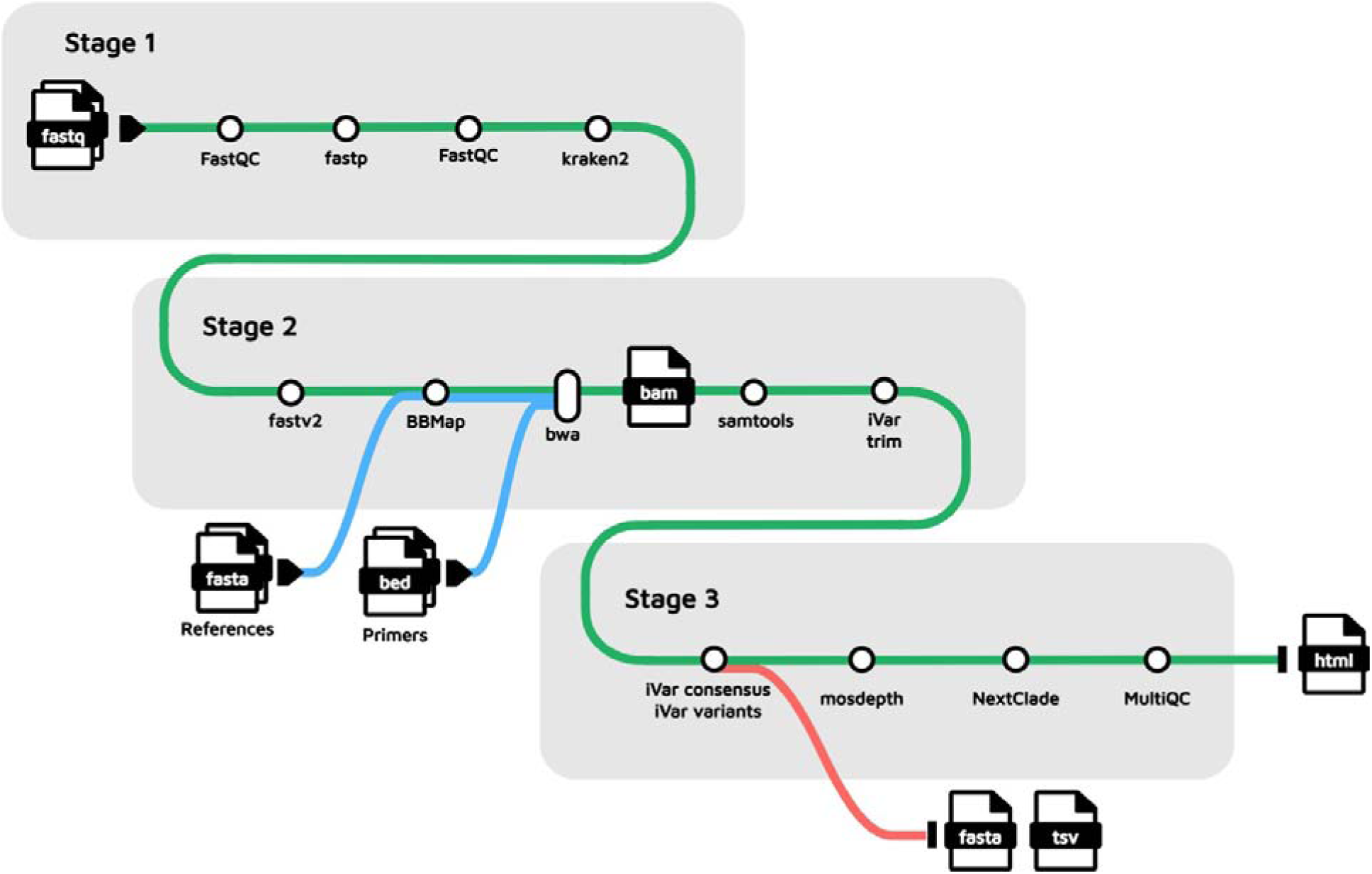
A custom, in-house bioinformatics pipeline was used to detect, subtype, and generate high-quality consensus sequences for RSV-A and RSV-B.

**Table 1.** List of Version-Controlled Software Tools used in GPHL-RSV-PIPE.

| Tool Description | Tool | Version | Reference |
| --- | --- | --- | --- |
| Read quality control | FastQC | 0.12.1 | Andrews, 2010 |
| Adapter trimming | Fastp | 0.23.4 | Chen, 2025 |
| Human host reads removal | Kraken2 | 2.1.3 | Wood, 2019 |
| Kmer detection | Fastv2 | 2.0 | Chen, 2021 |
| Reference selection | BBMap | 39.16 | Bushnell, 2014 |
| Read alignment | BWA | 0.7.17-6 | Li, 2013 |
| Sort and index BAM files | Samtools | 1.17 | Danecek et al., 2021 |
| Trimming, variant calling,<br>and consensus generation | iVar | 1.4.2 | Grubaugh et al.,<br>2019 |
| Coverage metrics | Mosdepth | 0.3.8 | Pedersen & Quinlan,<br>2018 |
| Phylogenetic analysis and<br>lineage assignment | Nextclade Web | 3.18.0 | Aksamentov et al.,<br>2021 |
| Statistics Aggregator | MultiQC | 1.25 | Ewels et al., 2016 |
| Data Visualization | R | 4.5.2 | R Core Team, 2025 |

## RESULTS

### Performance Comparison of Custom and ARTIC RSV Amplicon Sequencing Panels

Performance of a custom PrimalScheme-designed primer set [19] was evaluated against the established ARTIC RSV panel [16]. Commonly used high-quality RSV-A and RSV-B reference genomes were aligned and used as input for primer design. The custom design showed an insert overlap size of 75 nucleotides and amplicon length range of 750-800 bp for RSV-A and 650-700 bp for RSV-B yielded predicted genome coverages of 98% for both subtypes. This resulted in 27 primer pairs for RSV-A and 33 primer pairs for RSV-B. We evaluated primer specificity using the NCBI BLAST tool [22]. No matches to human genomic sequences were identified, confirming that host DNA would not be amplified and supporting the specificity of the primers for RSV sequencing. Additionally, the primers demonstrated 100% query coverage and 100% sequence identity to human RSV sequences in the NCBI nucleotide database. The ARTIC RSV panel comprises 50 primer pairs for each RSV-A and RSV-B, with amplicon sizes of approximately 400 bp that covered the full-genome [16]. For both primer sets, RSV-A and RSV-B primers were divided into odd and even pools for multiplex PCR.

To compare performance between the ARTIC RSV panel and the custom designed panel, the same set of 46 RSV-A and 63 RSV-B samples (109) were sequenced using both approaches. Mean sequencing depth, median depth per amplicon, and percent genomic coverage were evaluated. Overall, 78% (85/109) of specimens demonstrated higher mean depth and genome coverage when sequenced with the ARTIC RSV panel, compared with 22% (24/109) using the custom-designed panel. Seventy-two of 85 specimens showed greater than a 1% increase in genomic coverage with the ARTIC panel. Comparison of the median depth of coverage per amplicon across all specimens showed that amplicon dropouts were more pronounced with the custom-designed panel, particularly among RSV-B specimens, where consistent dropouts were observed between genomic positions 8,300 and 8,800 (Figures S1 and S2). Given the consistently higher sequencing depth, improved genomic coverage, and fewer amplicon dropouts (Figure S2), the ARTIC RSV primer set was selected for validation and implementation at GPHL.

### Analytical Performance of RSV Whole-Genome Sequencing Relative to PCR

Thermo Fisher TaqMan™ Gene Expression Capillary assay, a real-time PCR assay included in the Thermo Fisher TrueMark Respiratory Panel, was used as comparator method. This assay detects and characterizes 41 viral, bacterial, and fungal targets, including respiratory viruses such as RSV-A, RSV-B, SARS-CoV-2, Influenza A, and Influenza B. To assess the analytical validity of the WGS assay, performance metrices, including sensitivity, specificity, accuracy, positive predictive value (PPV), and negative predictive value (NPV), were evaluated using a sample set consisting of RSV-A, RSV-B, and PCR-negative clinical specimens tested by both PCR and WGS assays. Correlation analysis between the two assays demonstrated a strong linear relationship between WGS mean sequencing depth and PCR C_q_ value for both RSV-A and RSV-B (Figure 2).

**Figure 2.**
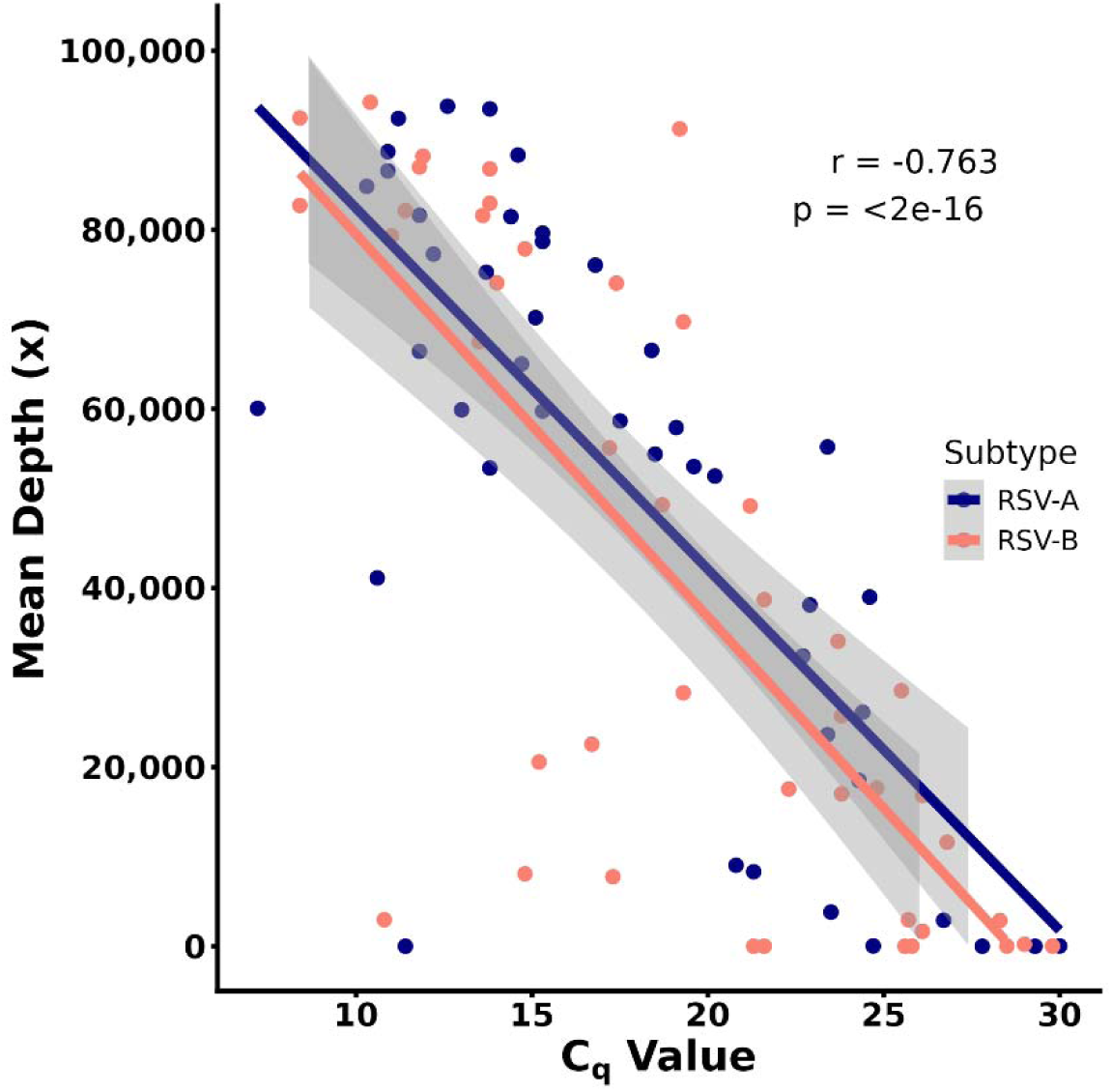
Correlation plot between mean WGS depth and C_q_ values from the comparator method, the Thermo Fisher TrueMark Respiratory Panel 2.0 TaqMan™ Array Card Assay, which includes a pre-amplification step prior to PCR. Although these are not true C_q_ values, a Pearson’s correlation coefficient of -0.76 indicates a strong negative correlation between C_q_ and sequencing depth.

WGS assay validation acceptance criteria for the clinical specimens required genomic coverage of at least 90%, with a mean depth of at least 100X. For RSV-A, 73.5% (75/102) of sequences met these criteria, with sequencing depth ranging from 16,265x to 83,811x (median, 53,433x; mean, 51,076x), and genomic coverage between 90.1% and 99.1% (median, 97.5%; mean, 96.6%), For RSV-B, 67.9% (76/112) sequences passed, with depth ranging from 6,386x to 106,560x (median, 49,699x; mean, 46,945x) and genomic coverage between 91.0% and 99.7% (median, 98.3%; mean, 97.6%) (Figure 3; Table S1). This indicates that sequencing depth was slightly higher for RSV-A genomes than for RSV-B, whereas genomic coverage was slightly lower for RSV-A. The lower passing rate for some specimens may be attributed to amplicon dropouts (Figure S2). Overall, the WGS assay demonstrated strong performance, with RSV-A showing an accuracy of 92.8%, sensitivity of 96.2% and specificity of 87.2% (Table 2), while RSV-B exhibited an accuracy of 93.2%, sensitivity of 97.4% and specificity of 84.6% (Table 2).

**Figure 3.**
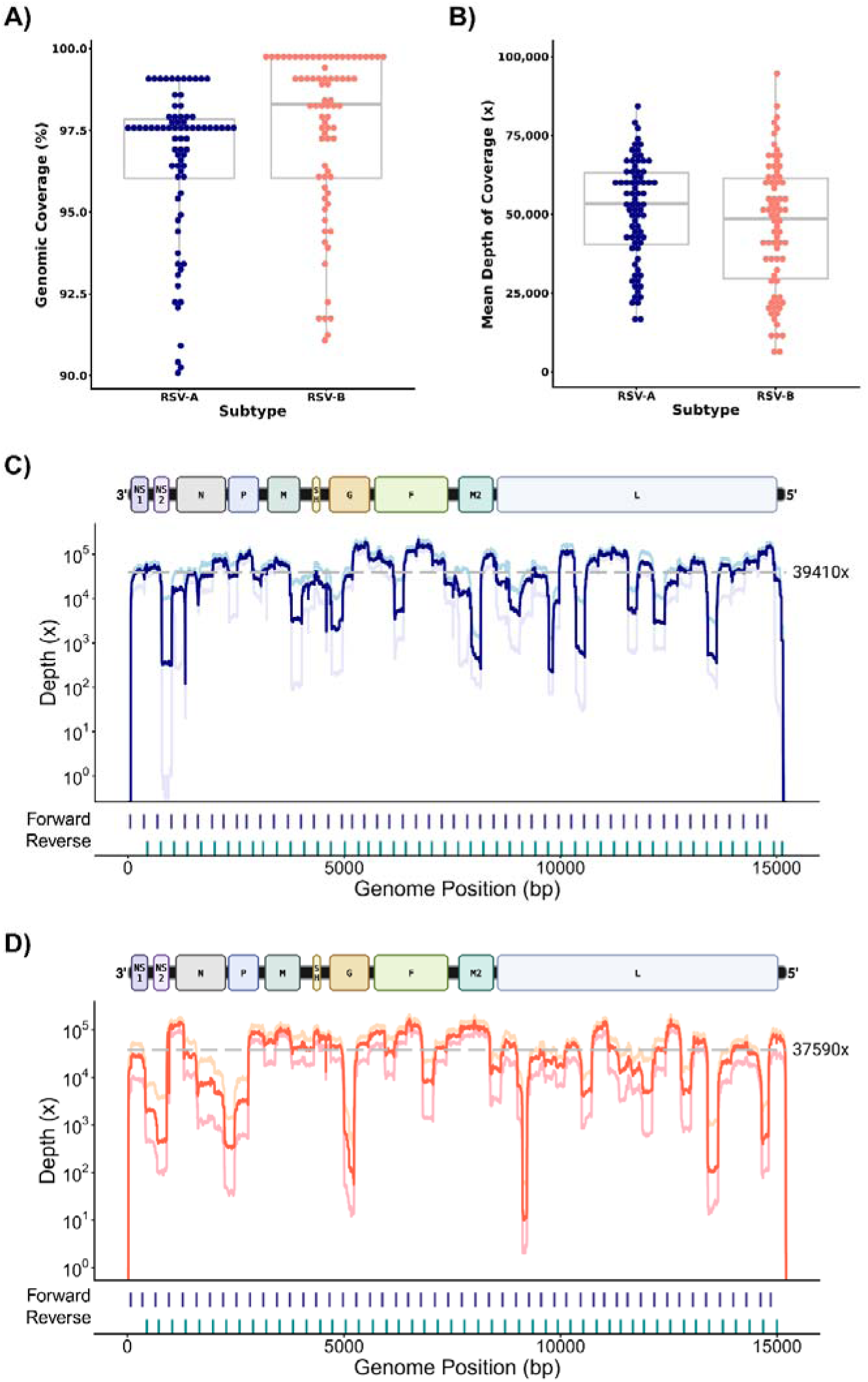
**A)** Genomic coverage and **B)** mean depth of coverage plots of high-quality RSV-A (75) samples and RSV-B samples (76). The boxplot values indicate the overall median percent coverage and depth: 98% and 53,434x for RSV-A, and 98% and 48,585x for RSV-B. **C)** Genomic coverage map for RSV-A; (Genomic coordinates: NS1 70-489, NS2 599-973, N 1111-2286, P 2318-3043, M 3226-3996, SH 4266-4460, G 4652-5617, F 5697-7421, M2-1 7640-8224, M2-2 8199-8465, L 8532-15029). **D)** Genomic coverage map for RSV-B; (Genomic coordinates: NS1 57-475, NS2 584-958, N 1097-2272, P 2305-3030, M 3154-3990, SH 4259-4456, G 4645-5578, F 5676-7400, M2-1 7627-8214, M2-2 8180-8452, L 8518-15018), showing all passing samples and primer pair positions. The dotted line indicates the median depth of coverage.

**Table 2.** Evaluation of the whole-genome sequencing assay with the comparator method, the Thermo Fisher TrueMark Respiratory Panel PCR assay, for RSV-A (A) and RSV-B (B). The number of PCR-positive samples that met the WGS validation acceptance criteria was 75 for RSV-A and 76 for RSV-B. Validation metrics were similar between RSV-A (**C**) and RSV-B (**D**), with all metrics exceeding 90%, except for specificity.

**A)**
| **PCR Result** | **WGS Positive** | **WGS Negative** |
| --- | --- | --- |
| Positive | 75 | 6 |
| Negative | 3 | 41 |

**B)**
| **PCR Result** | **WGS Positive** | **WGS Negative** |
| --- | --- | --- |
| Positive | 76 | 6 |
| Negative | 2 | 33 |

**C)**
| **Sensitivity** | **Specificity** | **Accuracy** | **PPV** | **NPV** |
| --- | --- | --- | --- | --- |
| 96.15 | 87.23 | 92.80 | 92.59 | 93.18 |

**D)**
| **Sensitivity** | **Specificity** | **Accuracy** | **PPV** | **NPV** |
| --- | --- | --- | --- | --- |
| 97.44 | 84.62 | 93.16 | 92.68 | 94.29 |

### Determination of the Lower Limit of Detection for RSV-A and RSV-B

The lower limit of detection (LOD) for RSV-A and RSV-B was determined using serial dilutions of reference material with known concentrations as Tissue Culture Infectious Dose 50 (TCID_50_/mL). For RSV-A, dilutions ranged from 280,000 to 0.35 TCID_50_/mL, while RSV-B dilutions ranged from 89,000 to 0.89 TCID_50_/mL (Table S2). Estimated LOD for RSV-A and RSV-B were calculated using probit regression analysis and were determined to be 4.4 TCID_50_/mL and 18.6 TCID_50_/mL, respectively (Figure 4). These estimates were subsequently confirmed using 20 replicate tests. For RSV-A, 18 of 20 replicates (90%) met the acceptance criteria, confirming an LOD of 4.4 TCID_50_/mL. For RSV-B, 19 of 20 replicates (95%) met the acceptance criteria, confirming an LOD of 18.6 TCID_50_/mL (Table S2).

**Figure 4.**
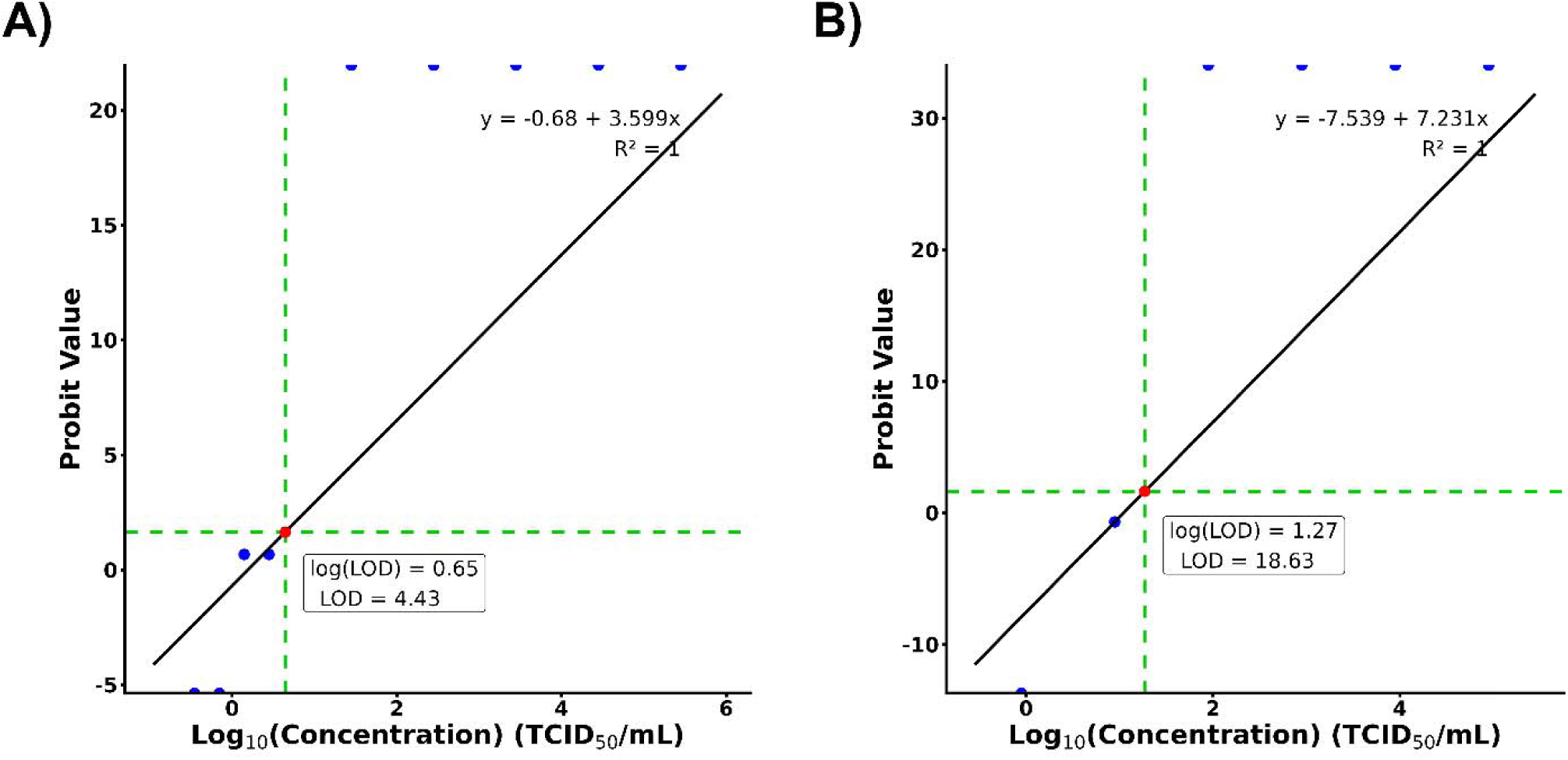
Probit regression analysis estimating the lower limit of detection as 4.4 TCID_50_/mL for (A) RSV-A and 18.6 TCID_50_/mL for (B) RSV-B. Data points at the top of each panel represent infinitive values.

### Performance of RSV WGS Assay with Mixed-Ratio RSV Specimens

A mix of RSV-A and RSV-B reference materials at varying ratios were used to evaluate the assay’s ability to detect multiple targets without interference in cases of co-infection. Ratios of equimolar concentrations of RSV-A to RSV-B included 100:0, 80:20, 60:40, 50:50, 40:60, 20:80, and 0:100. While the 100% mixes were run in singlicate, all other mixtures were run in triplicate. The 100% mixes correctly yielded positive detection for either RSV-A or RSV-B. For the mixed ratios, positive k-mer detection hits were observed for both RSV-A and RSV-B; during the alignment step, 93% (14/15) of replicates had most mapped reads aligned to the correct reference genome (Table S3).

### Specificity Assessment of RSV WGS Assay Against Other Respiratory Viruses

A set of 31 non-RSV samples, which tested positive for SARS-CoV-2 (n = 8), other coronaviruses (3), rhinovirus (5), mixed human parainfluenza virus 1 and 2 (1), human metapneumovirus (1), adenovirus (1), influenza A virus (8), and influenza B virus (4) was used to evaluate the specificity and cross-reactivity of the assay. The cutoff for the fastv k-mer detection tool was set at a minimum of 10% k-mer coverage or 500x k-mer depth to be considered positive. None of the 31 non-RSV samples produced positive detection hits for RSV-A and RSV-B, and no genome assemblies were generated (Table S4).

### Intra- and Inter-Run Performance of the WGS Assay

Repeatability (intra-run precision) and reproducibility (inter-run precision) were evaluated. For repeatability, 16 samples were run in duplicate by a single operator. After excluding regions with low-quality reads that resulted in ambiguous base calls (“N”), the consensus genomes for each sample pair showed nearly 100% identity. Fourteen of 16 specimens had no more than three pairwise nucleotide differences (Table S5). For reproducibility, three operators processed the same sample set on one Illumina NextSeq 1000. In addition, one operator ran the same set on one NextSeq 2000 and two NextSeq 1000 instruments to assess reproducibility across operators and sequencing instruments. Among positive specimens, percentage differences in genomic coverage between operators were less than 5%. The percentage coefficient of variation (%CV) across replicates was within an acceptable range of 10-20% for mean depth and less than 5% for genomic coverage. Furthermore, after excluding regions of ambiguous bases (“N”), these consensus genomes demonstrated nearly 100% pairwise identity (Tables S6 and S7) with 0-6 pairwise nucleotide differences.

### Phylogenetic Tree Construction and Clade Diversity

Maximum likelihood phylogenetic trees were constructed using 75 RSV-A and 76 RSV-B consensus sequences that met validation acceptance criteria, employing Nextstrain’s Augur and Auspice tools [38, 39] (Figure 5). The same lineages were formed with a cluster. For RSV-A, the predominant lineages were A.D.3.1 (n=26) and A.D.5.2 (21), together accounting for 63% (47/75) of the validated specimens. The remaining 37% comprised lineages A.D.1.5 (10), A.D.1.6 (4), A.D.1.8 (1), A.D.1.9 (1), A.D.2.1 (5), A.D.3 (2), A.D.3.2 (2), A.D.3.9 (2), and A.D.5.1 (1). For RSV-B, the dominant lineage was B.D.E.1, representing 91% (69/76) of the validated specimens. The remaining specimens belonged to clades B.D.E.1.2 (3), B.D.E.1.4 (1), B.D.E.1.8 (1), B.D.4.1.1 (1) and B.D.E.5 (1). Overall, these analyses demonstrated greater lineage diversity among RSV-A specimens compared to RSV-B.

**Figure 5.**
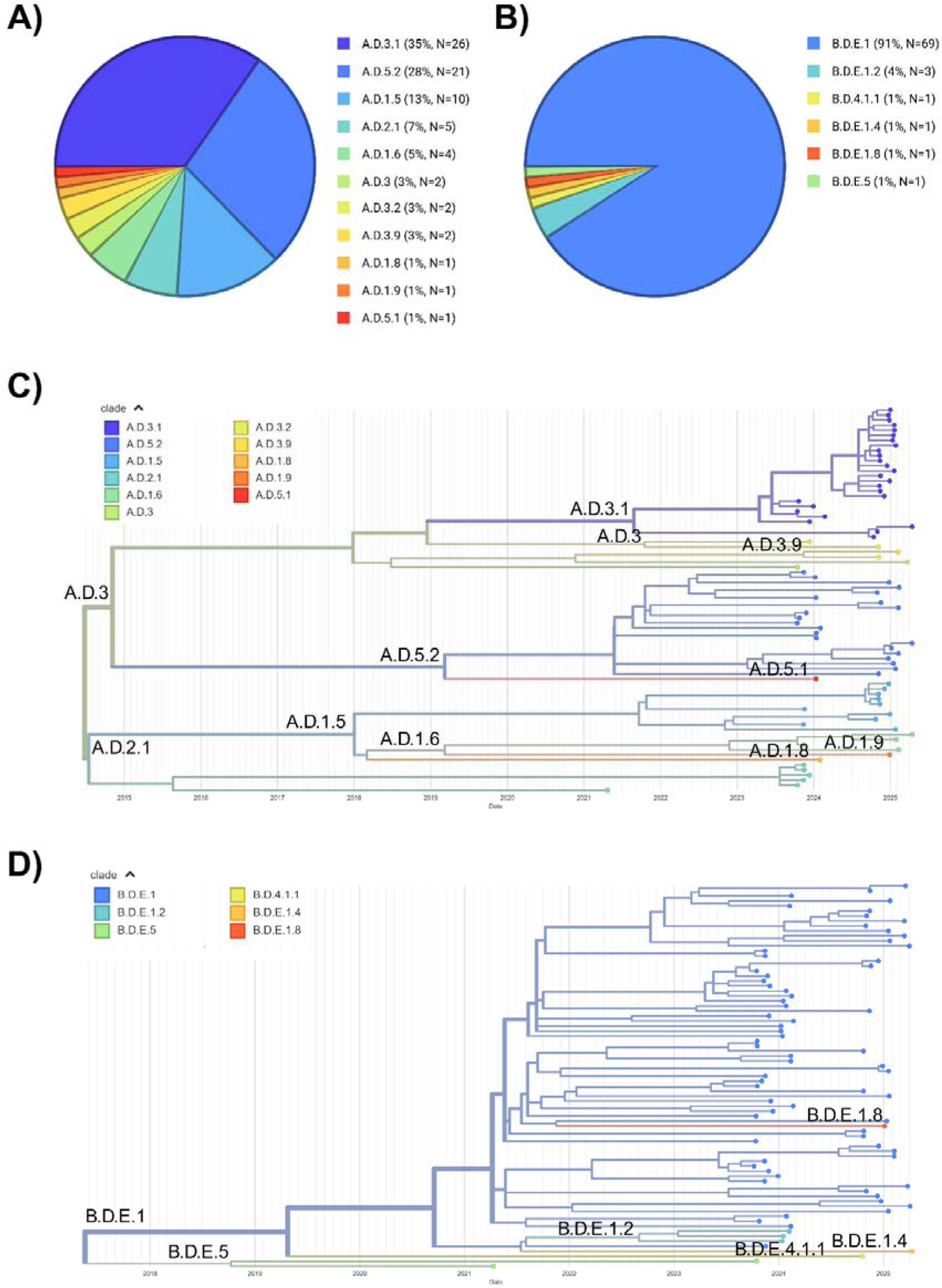
Lineage distribution and phylogenetic analysis of RSV-A and RSV-B. (A &. **B)** Pie charts show the distribution of clades in the sample sets. For RSV-A, the dominant clades were A.D.3.1 (n = 26) and A.D.5.2 (n = 21), together comprising 63% (47/75) of the passing samples. For RSV-B, the dominant clade is B.D.E.1 (n = 69), representing 91% (69/76) of passing samples. The maximum likelihood phylogenetic trees of (C) RSV-A and (D) RSV-B genomes generated in Nextclade.

## DISCUSSION

This investigation aimed to develop and validate a short-read, amplicon-based WGS assay at GPHL and to use an in-house–developed, reproducible bioinformatics pipeline to analyze two RSV subtypes in Georgia. This approach allows for the detection and subtyping of RSV, support lineage assignment, and facilitates the generation of high-quality consensus genome sequences. The initiative strengthens sequencing capacity in Georgia, improves understanding of circulating RSV strains, and supports timely public health action.

RSV WGS assay was developed by adapting the ARTIC RSV primer set as described [9–10, 16]. The method employs a tiled, amplicon-based multiplexed approach using the Illumina COVIDSeq Test kit. This protocol effectively generated high-quality consensus genomes for both RSV-A and RSV-B, achieving a median depth greater than 45,000x and median genomic coverage of 98%. The assay demonstrated high accuracy (>90%) and strong analytical sensitivity, with a LOD. Although specificity was below 90% when compared to the reference PCR method, the WGS assay remained highly specific, as no genome assembly or detection occurred in non-RSV samples. Cross-reactivity and mixed-sample evaluation of RSV-A and RSV-B sequence data confirmed that the in-house bioinformatics pipeline can accurately process co-infected samples. Replicate testing of the same samples within a single sequencing run, as well as across different operators and sequencing instruments, demonstrated high intra-run precision (repeatability), and inter-run precision (reproducibility). Multiple sequence alignments of replicate genomes showed near 100% concordance, with only minimal nucleotide differences observed.

Real-Time PCR-based assays are ‘gold-standard’ comparator methods for sequencing validations, as they provide ground-truth data for both specimen detection and copy number quantification [40–43]. The Thermo Fisher TrueMark Respiratory Panel 2.0 TaqMan™ Array Card Assay (cat# A49047; RVP) is designed to detect 41 known respiratory tract microbes, including RSV. This assay includes a pre-amplification step prior to PCR to enhance the signal of target sequences, which results in lower cycle threshold values for positive samples. Rather than reporting traditional C_t_, the RVP assay provides inflated C_q_ values. These values can reliably indicate the presence of a viral pathogen but cannot be used for true quantitative comparisons. Nonetheless, as noted previously, a strong linear correlation between the RVP C_q_ value and mean WGS depth of coverage provides general context that mean depth decreases as RVP C_q_ increases (Figure 2).

We encountered a few challenges during the validation process. First, the WGS assay was not compared to another sequencing-based assay, which posed challenges because PCR and WGS assays inherently differ in performance characteristics. PCR assays, such as the comparator method used in this study, are typically more sensitive and analytically optimized for targeted detection. In our analysis, three samples that tested positive for RSV-B by PCR were identified as RSV-A by WGS. Conversely, two samples that were positive for RSV-A by PCR were identified as RSV-B by WGS. These discrepancies affected specificity, as PCR and WGS assigned opposite subtypes to the same samples. One possible explanation is that mutations were present in the genomic regions targeted by the PCR assay, leading to misclassification of RSV subtype. However, the specific gene regions amplified by the RVP assay are not publicly available, limiting further investigation into this possibility. This highlights the challenges associated with comparing assays based on fundamentally different detection principles. Secondly, as previously mentioned, the comparator method, the RVP PCR assay, includes a pre-amplification step and therefore does not report a true C_q_ value. Consequently, we were unable to determine the viral load directly from the RVP assay. Instead, we relied on commercially available cultured specimens with predetermined concentrations to calculate the LOD. These concentrations were reported in TCID_50_/mL, a unit associated with cell culture, which is less common than copies/mL and can be challenging to translate for molecular applications. The third caveat involved amplicon dropouts due to mutations in primer-binding regions, specifically within the largely variable G gene of both RSV-A and RSV-B primer sets. Because RSV evolves rapidly, such dropouts are expected. To address this, we plan to routinely update primers as needed to capture emerging variants and maintain high sequencing depth and coverage.

The validated WGS assay, together with the optimized and reproducible bioinformatics pipeline, demonstrated that it is a robust, scalable, and cost-effective platform for routine near-real-time RSV genomic surveillance in Georgia. Further implementation of this WGS assay enhances Georgia’s ability to monitor viral evolution, identify emerging and re-emerging variants, detect transmission patterns, and inform prevention and response strategies, thereby supporting national public health efforts. The RSV WGS assay developed at GPHL serves as a robust and scalable model for public health laboratories seeking to enhance viral surveillance. Strengthening sequencing capacity in public health laboratories facilitates genomic epidemiology investigations and enables timely, data-driven decision-making.

## DATA AVAILABILITY

The raw reads for all RSV-A and RSV-B specimens were submitted to NCBI SRA under the BioProject number PRJNA1400014. The assembled genomes were submitted to GISAID database (Table S8).

## ETHICAL STATEMENT

The clinical specimen used in this study was received per Georgia Department of Public Health (DPH) - Institutional Biosafety Committee (IBC) guidelines. DPH IRB or Ethical Committee determined the laboratory diagnostics and genomic surveillance activities were for non-research public health surveillance and had no ethical concerns.

## ACKNOWLEDGMENTS

We acknowledge the Pathogen Genomics Center of Excellence (PGCOE) Network partner, the Minnesota Department of Health Laboratory, for sharing the initial sequencing protocol for RSV clinical samples. The authors gratefully acknowledge Melissa Tobin-D’Angelo, Director of the GDPH Emerging Infections Program (EIP), for providing some of the clinical specimens used in this assay validation. KS and JM thank the Association of Public Health Laboratories for supporting them through the APHL Bioinformatics Fellowship.

## SUPPLEMENTAL FIGURES LEGEND

**Figure S1.** Heatmap of (**A**) RSV-A (**B**) RSV-B showing the median depth of coverage per amplicon (x-axis) for samples meeting validation acceptance criteria (y-axis) using the PrimalScheme designed custom primers.

**Figure S2.** Heatmaps of RSV-A (**A**) and RSV-B (**B**) showing the median depth of coverage per amplicon (x-axis) across samples that met validation acceptance criteria (y-axis) using ARTIC primers.

## SUPPLEMENTAL TABLES LEGEND

**Table S1**. A total of 102 RSV-A and 112 RSV-B clinical samples were sequenced. 75 out of 102 RSV-A samples passed while 75 out of 112 RSV-B samples passed the validation acceptance criteria. Whole-genome sequencing was compared to the reference method, Thermo Fisher Respiratory Viral Panel PCR assay. Mean depth and percent genomic coverage were evaluated. *Please refer to the Table_S1 sheet in the Excel file*.

**Table S2**. The lower limit of detection (LOD) for RSV-A and RSV-B was determined by an initial test of finding the estimated LOD with serial dilutions (A). Replicates LOD confirmation for RSV-A (B) and RSV-B (C). *Please refer to the Table_S2 sheet in the Excel file*.

**Table S3**. Mixtures of RSV-A and RSV-B reference material were used to evaluate multiplexing and ability to detect multiple targets with the WGS assay. All mixes passed validation acceptance criteria. 14/15 (93%) of replicates had majority of the mapped reads aligned to the correct target reference genome. *Please refer to the Table_S3 sheet in the Excel file*.

**Table S4**. A variety of 31 non-RSV samples was sequenced to evaluate specificity and cross-reactivity of the WGS assay as part of the validation. All samples did not have any positive kmer detection hits for both RSV-A and RSV-B. *Please refer to the Table_S4 sheet in the Excel file*.

**Table S5**. Pairwise nucleotide differences between duplicates of the same sample within the same sequencing run by one operator for intra-run precision (repeatability) among 16 high-quality RSV-A and RSV-B samples that passed validation acceptance criteria. Each pair of consensus sequences were nearly 100% identical with 14 out of 16 samples had no more than 3 nucleotide differences. *Please refer to the Table_S5 sheet in the Excel file*.

**Table S6**. Pairwise nucleotide differences between 3 operators for inter-run precision (reproducibility) among 53 high-quality RSV-A and RSV-B samples that passed validation acceptance criteria. Each pair of consensus sequences were nearly 100% identical with 0 to 5 nucleotide differences. *Please refer to the Table_S6 sheet in the Excel file*.

**Table S7**. Pairwise nucleotide differences between 3 separate sequencers (2 NextSeq 1000s, 1 NextSeq 2000) for inter-run precision (reproducibility) among 57 high-quality RSV-A and RSV-B samples that passed validation acceptance criteria. Each pair of consensus sequences were nearly 100% identical with 0 to 4 nucleotide differences. *Please refer to the Table_S7 sheet in the Excel file*.

**Table S8.** GISAID Accession IDs for 151 submitted complete RSV genomes. *Please refer to the Table_S8 sheet in the Excel file*.

