## Supplemental Figures for "Development and Evaluation of an ARTIC-Based Amplicon Sequencing Assay for Whole-Genome Characterization of Respiratory Syncytial Virus"

**Figure S1.** Heatmap of (**A**) RSV-A (**B**) RSV-B showing the median depth of coverage per amplicon (x-axis) for samples meeting validation acceptance criteria (y-axis) using the PrimalScheme designed custom primers.

A

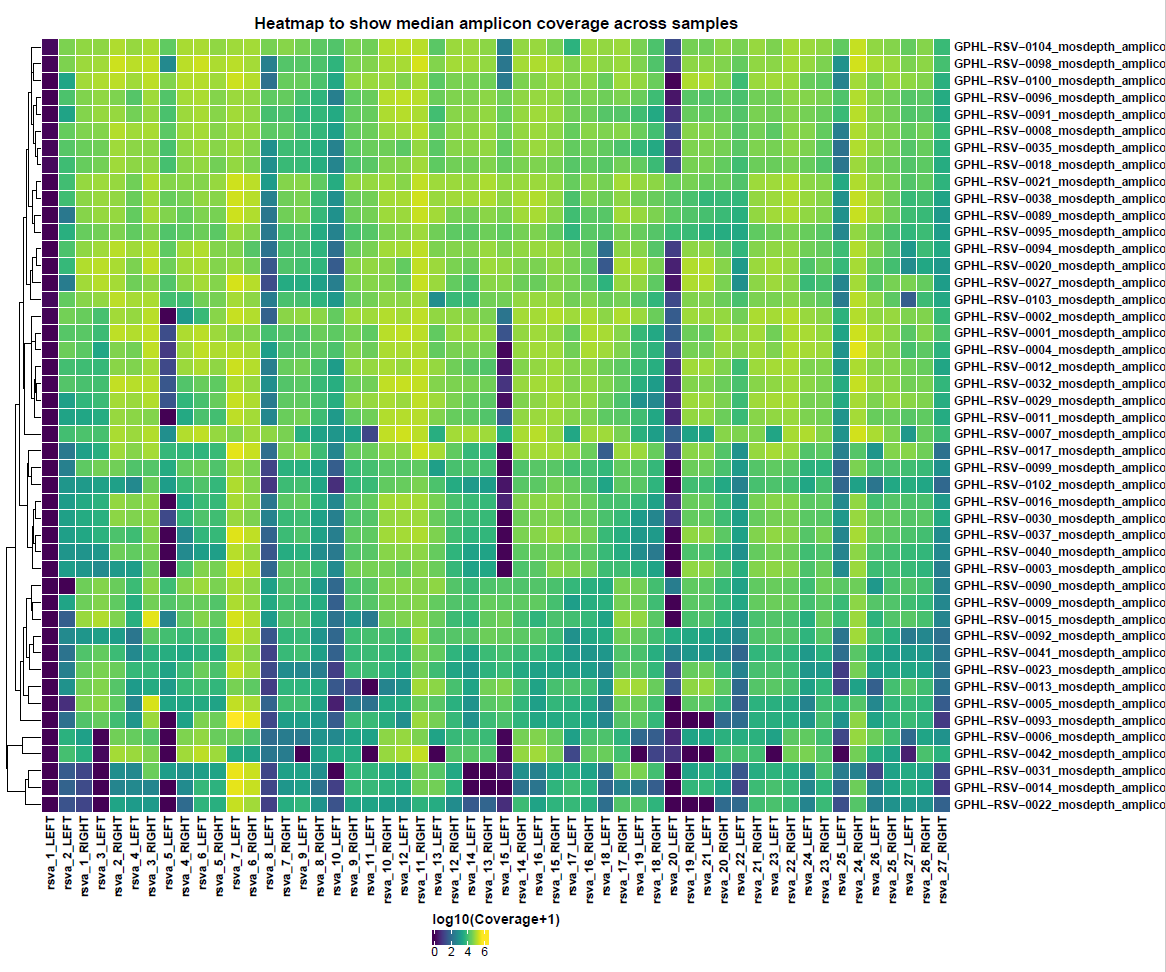

B

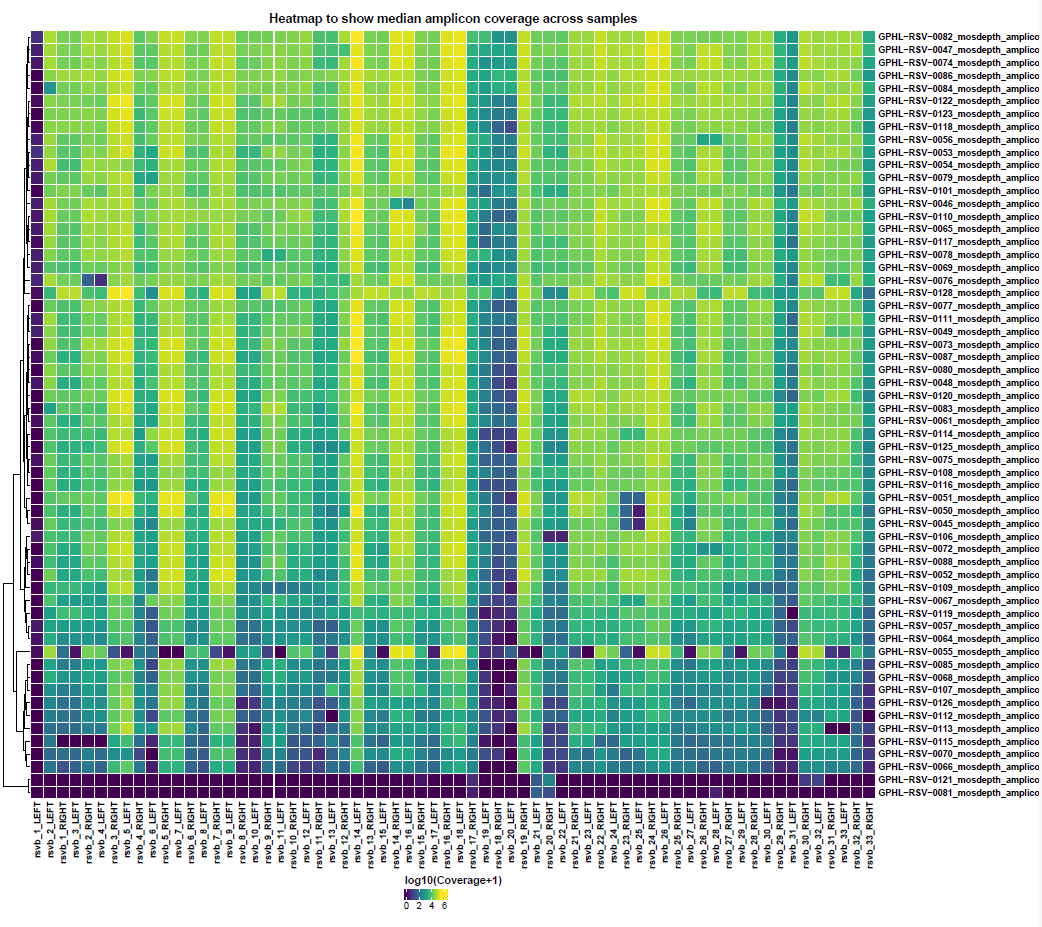

**Figure S2.** Heatmaps of RSV-A (**A**) and RSV-B (**B**) showing the median depth of coverage per amplicon (x-axis) across samples that met validation acceptance criteria (y-axis) using ARTIC primers.

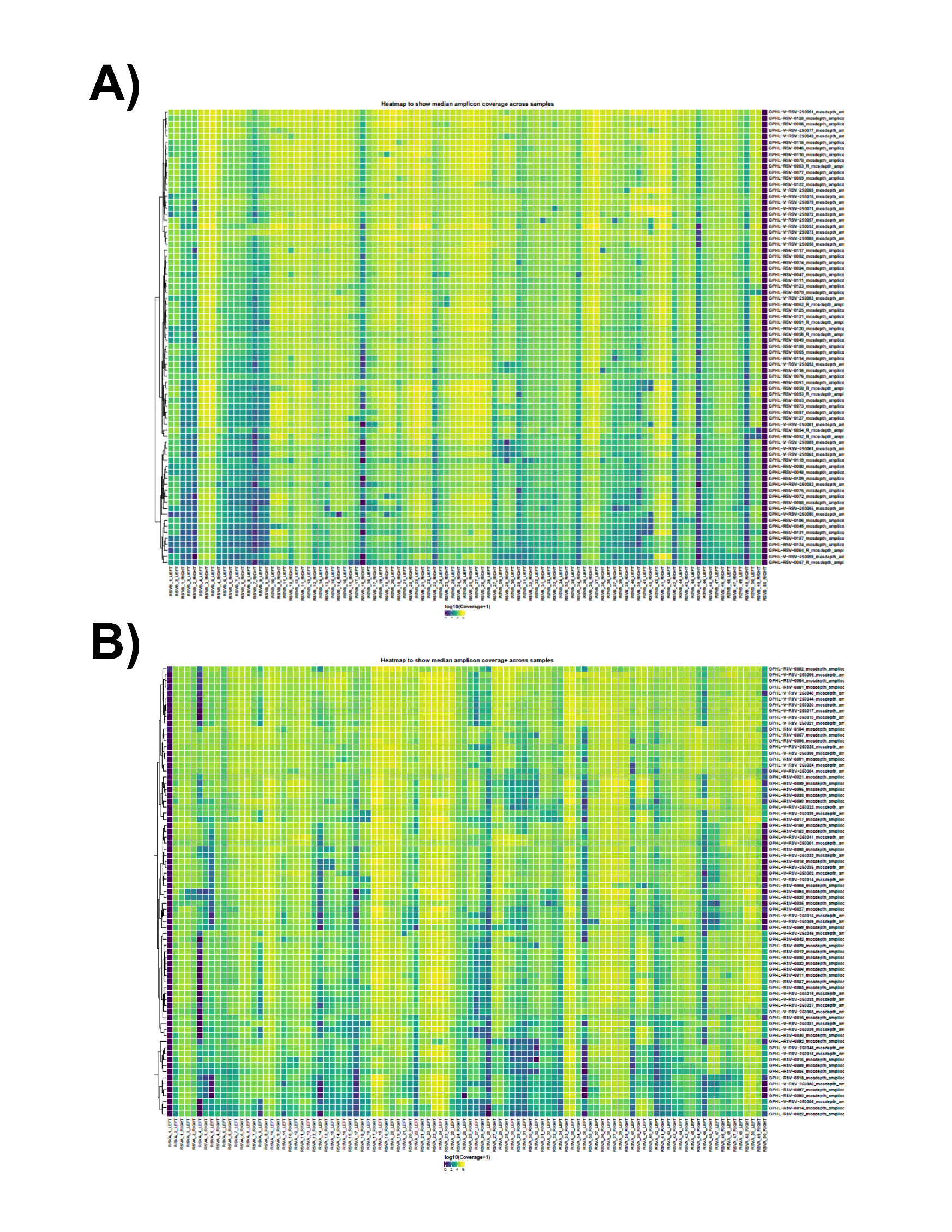
A

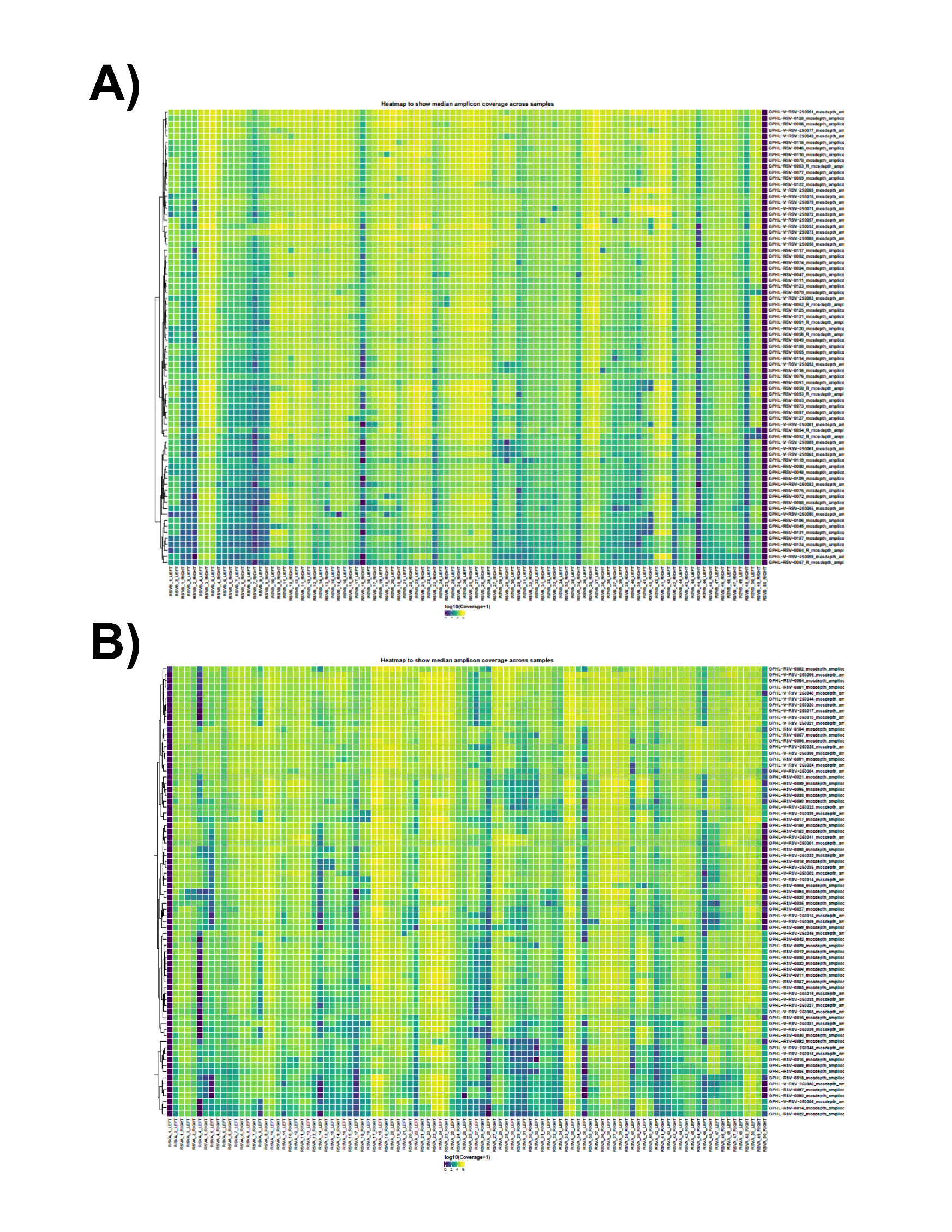
**B**
